# The production of anti-PF4 antibodies in anti-phospholipid antibody-positive patients is not affected by COVID-19 vaccination

**DOI:** 10.1101/2021.08.12.21261976

**Authors:** Paola A. Lonati, Caterina Bodio, Mariangela Scavone, Giuliana Martini, Elisa Pesce, Alessandra Bandera, Andrea Lombardi, Maria Gerosa, Franco Franceschini, Angela Tincani, Gianmarco Podda, Sergio Abrignani, Renata Grifantini, Marco Cattaneo, Maria Orietta Borghi, Pier Luigi Meroni

**Author notes:** **CORRESPONDENCE to** Prof. Pier Luigi Meroni, Experimental Laboratory of Immunological and Rheumatologic Researches, Istituto Auxologico Italiano, Milan, Italy. PAL and CB contributed equally to this paper. MOB and PLM shared the senior autorship.

## Abstract

Antibodies against cationic platelet chemokine, platelet factor 4 (PF4/CXCL4) have been described in heparin-induced thrombocytopenia (HIT) but also in patients positive for anti-phospholipid antibodies (aPL) even in the absence of heparin treatment and HIT-related clinical manifestations. Anti-PF4 antibodies have been recently described also in subjects who developed thrombosis with thrombocytopenia syndrome (TTS) in association with adenoviral vector-based, but not with mRNA-based COVID-19 vaccines.

We investigated whether COVID-19 vaccination affects the production of anti-PF4 immunoglobulins detectable by solid phase assay in aPL-positive patients and their ability to induce in vitro platelet activation. Anti-PF4 were found in 9/126 aPL-positive patients, 4/50 COVID-19, 9/49 other infections and 1/50 aPL-negative systemic lupus erythematosus patients. Clinical manifestations of TTS were not observed in any aPL patient positive for anti-PF4, whose sera failed to cause platelet aggregations. The administration of COVID-19 vaccines did not affect the production of anti-PF4 immunoglobulins or their ability to cause platelet aggregation in 44 aPL-positive patients tested before and after vaccination. In conclusion, heparin treatment-independent anti-PF4 antibodies can be found in aPL-positive patients and asymptomatic carriers, but their presence, titer as well as in vitro effect on platelet activation are not affected by COVID-19 vaccination.

## INTRODUCTION

Antibodies against cationic platelet chemokine, Platelet Factor 4 (PF4/CXCL4) have been reported in heparin-induced thrombocytopenia (HIT) but also in patients positive for anti-phospholipid antibodies (aPL) even in the absence of treatment with heparin and HIT-related clinical manifestations.[1-7] Despite the heterogeneity of published data, the majority of the studies reported the presence of anti-PF4 antibodies in up to 21% of aPL-positive samples, their heparin-dependent binding as in HIT but at lower titer and no effect on platelet activation.

Anti-PF4 antibodies have also been recently described in COVID-19 vaccine-associated thrombosis with thrombocytopenia syndrome (TTS).[8-11] Although the hypothesis of a molecular mimicry between self-autoantigens and SARS-CoV-2 spike (S) protein is still debated, the active immunization with S protein was suggested to be responsible for the antibody response against PF4 as well.[12, 13] Accordingly, the issue of a potential danger of COVID-19 vaccination in aPL-positive patients was raised because of their thrombophilic state and the possible occurrence of anti-PF4 antibodies in some of them. With this in mind, we investigated whether COVID-19 vaccination affects the production of anti-PF4 antibodies in aPL-positive patients and their functional ability to induce in vitro platelet activation.

## METHODS

### Patients

We investigated 126 aPL-positive samples including 71 primary anti-phospholipid syndrome (PAPS), 37 aPL-positive asymptomatic carriers, 18 APS associated with systemic autoimmune rheumatic disorders (SAPS). The diagnoses were made as previously described;[14] all samples displayed double or triple positivity for the APS laboratory classification criteria,[14] and medium/high titers of anti-cardiolipin and anti-beta2 glycoprotein I IgG/IgM.

As control groups the following samples were also tested: 50 COVID-19 patients with moderate disease as previously reported,[15] 49 individuals with non-COVID-19 infections (9 Epstein-Barr virus, 2 hepatitis C virus, 14 rubella virus, 14 cytomegalovirus, 10 syphilis) and 50 aPL-negative systemic lupus erythematosus (SLE) patients.[16]

Nineteen PAPS, 12 aPL-positive asymptomatic carriers, and 13 SAPS were tested before and after COVID-19 vaccination (38 with Comirnaty, 2 with Spikevax, 3 with Vaxzevria, and 1 with Sputnik vaccine). Additional 2 PAPS patients were tested before and after full blown COVID-19 and positivity for SARS-CoV-2 nasopharyngeal swab. One-hundred fifty healthcare workers were also enrolled and serum samples were collected before and after vaccination by Comirnaty (100) or Vaxzevria (50).

Samples were collected before the second vaccine injection for both aPL-positive patients and healthcare workers (3 weeks after Comirnaty or Sputnik, 4 weeks after Spikevax and 12 weeks after Vaxzevria first injection, respectively).

The severe adverse side effects as defined by Polack et al.[17] or any clinical manifestation potentially correlated with the vaccination were also recorded for all the investigated patients or subjects.

The Ethics Committee at Istituto Auxologico Italiano approved the study (08-01-2021). All the patients/subjects gave their informed consent.

### Anti-PF4 detection

Anti-PF4 IgG/IgA/IgM were assessed by the polyspecific Lifecodes PF4 Enhanced ELISA (Immucor, Solihull, UK). The assay was performed according to the manufacturer’s instructions, including negative and positive controls and confirmatory inhibition of the reaction in the presence of high concentrations of heparin (100 U/ml). Anti-PF4 antibodies are detectable in 1.0-4.3% of normal healthy subjects (NHS), depending on the commercial kit used.[18] Because of this variability of results, we used our in-house cut-off value of 0.80 optical density (OD) units, which was calculated as the mean + 3 SD of the results obtained in 189 NHS. Samples with binding values > 0.80 OD were re-tested in the presence of heparin (100 IU/mL) and for their ability to induce platelet aggregation (Platelet Aggregation Test, PAT).

### Platelet Aggregation Test

PAT was performed using washed human platelet (WP) suspensions prepared as described [19] from ACD-anticoagulated blood from NHS and resuspended with Tyrode’s solution without added CaCl_2_, containing 0.01 U/mL Apyrase from potatoes (Sigma-Aldrich, Taufkirchen, Germany). Heat inactivated (56°C, 30 min) serum was added to WP with buffer or heparin (0.2 and 100 IU/mL) (Veracer; Medic Italia, Milan, Italy). Platelet aggregation was measured in the PAP-8E Platelet Aggregation Profiler (Bio/Data Corporation, Horsham, PA, USA) for 30 min.

## RESULTS

### Prevalence of anti-PF4 immunoglobulins in the study groups

Figure 1 shows the prevalence of anti-PF4 immunoglobulins in the study groups. Values of anti-PF4 immunoglobulins higher than the cut-off value of 0.80 OD were found in 9/126 (7%) aPL-positive samples, 1/50 (2%) aPL-negative SLE, 4/50 (8%) COVID-19 and 4/49 (18%) patients suffering from non-COVID-19 infectious diseases. The antibody binding in the presence of excess of heparin (100 IU/mL) was significantly inhibited in all positive samples. Moreover, the titers of anti-PF4 immunoglobulins were lower than those usually described in HIT patients and no HIT related clinical manifestations were recorded.[11]

**Figure 1.**
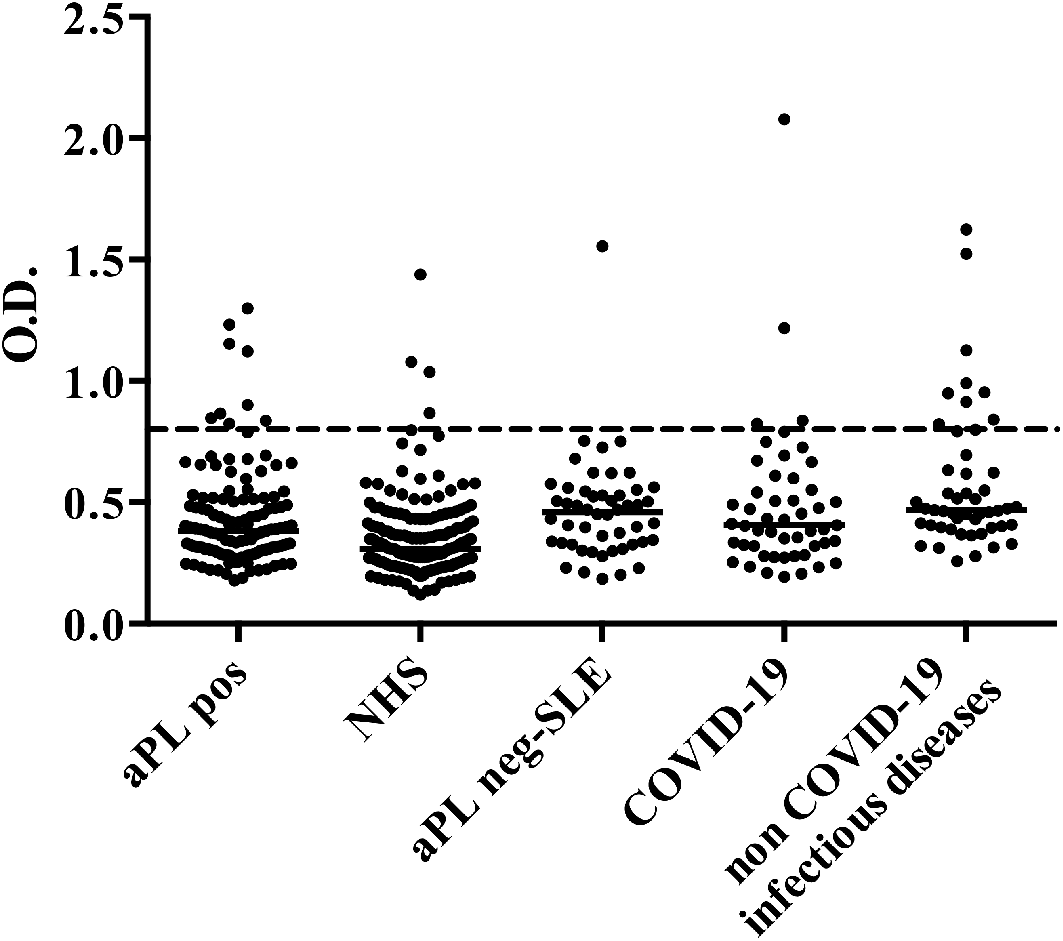
Binding activity of anti-PF4 immunoglobulins in the study groups. Anti-PF4 binding activity of serum samples from aPL-positive patients or carriers (aPL pos), normal healthy subjects (NHS), SLE patients negative for aPL (aPL neg-SLE), patients with COVID-19 or non-COVID-19 infections. Data are expressed as optical density (OD). Dashed line indicates the in-house cut-off value (0.80 OD).

### Variations of anti-PF4 immunoglobulin titers before (T0) and after (T1) COVID-19 vaccination

Figure 2A shows the variations of anti-PF4 antibody reactivity before and after Covid-19 vaccination in aPL-positive patients classified as PAPS, SAPS or asymptomatic aPL-positive carriers. No significant changes in antibody titers were found in all the investigated patients. Two additional patients with PAPS were tested before and one month after full-blown COVID-19 of moderate severity and did not show any modification in the titers (PAPS1: 0.476 OD and 0.423 OD, PAPS2: 0.443 OD and 0.788 OD before and after COVID-19, respectively).

**Figure 2.**
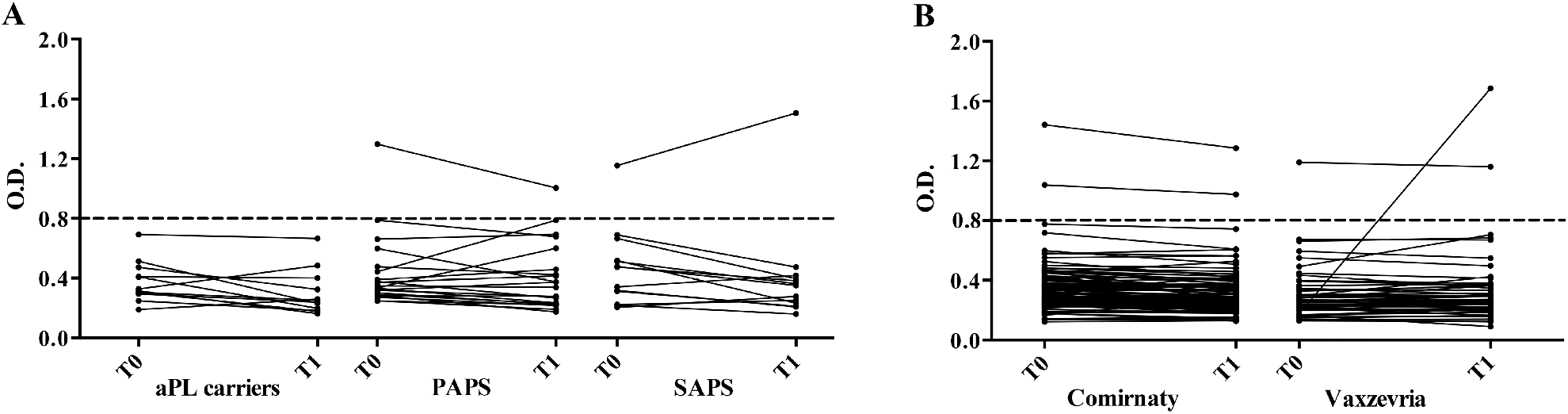
Effect of COVID-19 vaccination on anti-PF4 immunoglobulin titers. (A) Anti-PF4 OD values before (T0) and after (T1) Covid-19 vaccination in asymptomatic aPL-positive carriers, PAPS and SAPS patients. (B) Anti-PF4 OD values before (T0) and after (T1) Comirnaty or Vaxzevria vaccines in healthcare workers. Dashed lines indicate the in-house cut-off value (0.80 OD).

Anti-PF4 immunoglobulin titers in healthcare workers before and after COVID-19 vaccination are shown in the Figure 2B. Anti-PF4 immunoglobulins at low titers were found at the baseline in 2/100 (2%) Comirnaty and in 1/50 (2%) Vaxzevria vaccinated subjects. An increase in the antibody titer was found in one subject only (from 0.174 OD to 1.682 OD) with no TTS-related symptoms and normal platelet count. No clinical manifestations related to TTS were recorded for all the other subjects included in the study and none of the positive sera (OD values > 0.80) tested positive in the PAT.

## DISCUSSION

Anti-PF4 immunoglobulins at low titer are detectable in a minority of healthy subjects and in different pathological conditions. In particular, our data show a prevalence of anti-PF4 positivity in SLE similar to that of the largest studies published in the literature.[1-7] These autoantibodies have been defined as “false-positive tests for HIT” because they are not associated with HIT-related clinical manifestations and do not trigger in vitro platelet activation.[1-7] Nevertheless, most of them display an in vitro heparin-dependent binding activity in spite of no treatment with heparin.[1-7] We confirmed and extended this finding showing low titer, heparin-dependent and PAT-negative anti-PF4 antibodies in a large series of aPL-positive patients, as well as in aPL-negative SLE and infectious diseases.

In agreement with others and in contrast with the data reported by Pauzner et al.,[1] we found a low prevalence of anti-PF4 immunoglobulins in PAPS, SAPS and SLE.[2-7] The addition of an excess of heparin strongly inhibited antibody binding in the solid-phase assay in all the samples, suggesting that the autoantibodies were heparin-dependent. At variance with the antibodies detectable in HIT, anti-PF4 antibodies in aPL-positive patients were at medium-low titer and without any platelet activation effect, even in the presence of low heparin concentration (0.2 IU/mL).

COVID-19 vaccination with adenovirus-based vaccines may trigger TTS associated with the presence of high titer anti-PF4 antibodies, which may trigger in vitro platelet activation even in the absence of low concentrations of heparin.[8-11] Whether or not COVID-19 vaccination may increase the titer of pre-existing anti-PF4 antibodies in aPL-positive patients or induce the ability to activate platelets is an issue with clinical implications because of the prevalence of aPL positivity in a small but significant percentage of the general population.[20] Our data show that vaccination against COVID-19 cannot trigger the ex-novo production of anti-PF4 antibodies nor affect the titer of preexisting antibodies in a well characterized aPL-positive series. More importantly, vaccination does not induce the ability of these antibodies to cause platelet activation in vitro. Comparable data were found in a group of healthcare workers vaccinated with Comirnaty or Vaxzevria, with the exception of the increase of anti-PF4 immunoglobulins in one subject only without any clinical or laboratory manifestations of TTS.

In summary, anti-PF4 antibodies can be found in a small proportion of aPL-positive patients but with characteristics different from the antibodies detectable in HIT and TTS patients. COVID-19 vaccination is apparently safe in aPL-positive patients and does not trigger the production of TTS-associated autoantibodies, although larger series of patients vaccinated with adenoviral vector-based vaccines are needed to definitely support our conclusions.

## Data Availability

The datasets used and/or analyzed during the current study are available from the corresponding author on reasonable request.

## KEY MESSAGES

### What is already known about this subject?

- Anti-PF4 antibodies have been described both in HIT and in aPL-positive patients independently from heparin treatment and HIT-related clinical manifestations.
- Anti-PF4 antibodies have been detected after the administration of adenoviral vector-based, but not mRNA-based COVID-19 vaccines and have been associated with TTS.

### What does this study add?

- Low-titer, heparin-dependent and PAT-negative anti-PF4 antibodies have been found in a small proportion of aPL-positive patients, as well as in aPL-negative SLE and infectious diseases.
- COVID-19 vaccination neither affects the titer of preexisting anti-PF4 antibodies in aPL-positive patients nor induces the ability of these antibodies to activate platelets in vitro.

### How might this impact on clinical practice or future developments?

- Vaccines against COVID-19 are seemingly safe and unable to induce the clinical and laboratory TTS-associated manifestation in aPL-positive patients.

## COMPETING INTERESTS

There are no competing interests for any author.

## CONTRIBUTORSHIP

PLM and MOB conceived the study and wrote the first draft of the manuscript. PAL, CB, MS, GM, EP performed the assays. AB, AL, MG, FF, AT, PLM recruited the patients/healthy volunteers and collected the clinical records. GP, SA, RF, MC, MOB and PLM analyzed the results. All the Authors revised and approved the final version of the manuscript.

## ACKNWOLEDGMENTS

The Authors gratefully aknowledge Stefania Bertocchi, Paolo Semeraro, Cecilia Nalli, Laura Andreoli, Stefania Masneri (ASST Spedali Civili di Brescia, Brescia, Italy) and Alfredo Canè (Istituto Auxologico Italiano, IRCCS, Milan, Italy) who took part in the vaccine administration. The Authors are indebted to all the patients and healthy volunteers who participated in this study.

## FUNDING

The paper was supported in part by Ricerca Corrente 2020 and 2021 - Ministero della Salute, Italy - to PLM.

## PATIENTS AND PUBLIC INVOLVEMENT

Patients and public were not involved in the design of the study. During the initial phases of the study, we obtained feedback from the patients. The results of the study will be disseminated in lay versions by Istituto Auxologico Italiano public relations and communications department for the benefit of the public.

## AVAILABILITY OF DATA AND MATERIALS

The datasets used and/or analysed during the current study are available from the corresponding author on reasonable request.

## Notes

### Competing Interest Statement

The authors have declared no competing interest.

### Author Declarations

Ethics approval ID : 08-01-2021 Istituto Auxologico Italiano

